# Enhancing Breast Cancer Classification: A Few-Shot Meta-Learning Framework with DenseNet-121 for Improved Diagnosis

**DOI:** 10.1101/2024.10.04.24314684

**Authors:** Nidhi Upadhyay, Anuja Bhargava, Upasana Singh, Mohammed H. Alsharif, Ho-Shin Cho

## Abstract

Breast cancer is a significant health concern globally, requiring early and accurate detection to improve patient outcomes. However, manual detection of breast cancer from medical images is time-consuming and inaccurate. Accurate assessment of cancer stages is critical for effective treatment and post-diagnosis handling. The goal of this research is to develop a specialized meta-learning method for classifying breast cancer images, particularly effective when working with limited data. Traditional cancer stage classification methods often struggle with insufficient labeled data, but meta-learning addresses this challenge by rapidly adapting to new tasks with few examples. The proposed method begins with image segmentation to identify regions of interest in the medical images, followed by thorough feature extraction to capture essential data representations. The critical meta-training phase involves refining a classifier within a metric space, utilizing cosine distance and an adaptable scale parameter. During the meta-testing stage, the adapted classifier predicts cancer stages using minimal support samples, achieving approximately 96% accuracy. This approach shows significant promise for the medical field, providing practical solutions to enhance diagnostic processes and improve predictions for breast cancer detection and treatment.

## Introduction

Breast cancer detection presents a significant challenge due to the scarcity of comprehensive and diverse datasets. This scarcity hinders the development of accurate classification models, which rely heavily on ample data for effective training. The issue of limited data availability is common in medical imaging, stemming from privacy concerns, high costs of data acquisition, and the difficulty of obtaining labeled samples. Consequently, medical datasets are often small-scale, presenting a major hurdle for traditional machine learning and deep learning methods [1]. The challenge of few-shot classification, where the goal is to classify with minimal examples, becomes particularly pertinent in this context. Medical imaging, and specifically breast cancer datasets, often suffer from class imbalances and limited labeled samples, further complicating to the development of effective techniques. To address this, convolutional neural networks for image classification, transfer learning to improve detection accuracy, and support vector machines (SVMs) and random forests for effective classification. Meta-learning frameworks and few-shot learning approaches have been utilized to address the challenge of narrowed annotated data, while hybrid models and ensemble methods have combined multiple techniques to enhance performance. These contributions highlight the continuous improvement and innovation in area of breast cancer detection.

The author [15] employed transfer learning with YOLO models (YOLOv3, YOLOv5, YOLOv5-Transformer) to improve breast cancer detection in mammograms, utilizing CBIS-DDSM, INbreast, and a proprietary dataset. The small YOLOv5 model achieved the best performance with a mean Average Precision (mAP) of 0.621. Eigen-CAM was employed for model introspection, effectively reducing false negatives by highlighting suspicious regions, though it increased false positives. Despite challenging anomalies, the model demonstrated strong detection capabilities, and its outputs, when combined with Eigen-CAM saliency maps, require clinical evaluation, making it a reliable tool for supporting clinical decisions. The author [16] employed a hybrid approach integrating image and numerical data features for breast cancer (BC) detection. This approach utilized the U-NET model with transfer learning for precise image segmentation and combined classifiers like random forest (RF) and support vector machine (SVM). These methods were evaluated using datasets such as the Wisconsin Breast Cancer Dataset (WBCD), demonstrating improved detection accuracy and highlighting the efficacy of integrating diverse data modalities in BC diagnosis.

In [17], the author introduces a cutting-edge method aimed at enhancing breast cancer (BC) detection through automated systems. This method focuses on nuclei detection and tumor classification, incorporating contrast enhancement and image segmentation via Linear Scaling centered Canny Edge Detection (LS-CED). The process involves a Soft Plus-Max Region-centered Fully Convolutional Network (SPM-R-FCN) for precise nuclei identification, followed by tumor cell (TC) classification based on size and shape using an adaptive threshold function. The extracted features are then analyzed by the SDM-WHO-RNN classifier, which classifies cancer as benign, malignant, or normal, achieving a remarkable accuracy of 97.9% in simulations. In [18], another innovative approach for breast cancer detection is proposed, leveraging multi-wavelength interference (MWI) phase imaging and hyperspectral (HS) imaging. This method analyzes blue (446.6 nm) and red (632 nm) interference patterns using Fast Fourier (FF) transform to identify refractive index variations between tumor and normal tissue. The classification algorithm demonstrates 94% specificity and 90.9% sensitivity with ex-vivo breast samples. This approach, which uses unstained samples, holds potential for precise tumor excision and in-vivo detection using standard RGB cameras.

The author [19] presents a method for early breast cancer (BC) detection using Transfer Learning techniques, VGG, ResNet, MobileNetV2—integrated with LSTM to analyze Ultrasound Images (USIs). To balance features, SMOTE was employed. The VGG16 model achieved an F1 score of 99.0%, MCC and Kappa Coefficient of 98.9%, and an AUC of 1.0. Cross-validation yielded an average F1-score of 96%. Grad-CAM and LIME enhanced visualization and interpretability, while confidence intervals confirmed robustness. This approach outperformed six state-of-the-art TL models, proving its effectiveness. The author [20] addresses breast cancer detection using mammogram images analyzed with deep learning models. Utilizing four public databases, each with 986 mammograms across normal, benign, and malignant categories, the research employs VGG-11, Inception v3, and ResNet50. An ensemble method with a modified Gompertz function for fuzzy ranking integrates decision scores adaptively. This approach outperformed other methods, with the ResNet50 ensemble achieving a classification accuracy of 98.98%.

The author [21] enhances breast cancer detection in mammography images using advanced AI techniques, specifically combining YOLOv5 and Mask R-CNN models. YOLOv5 detects and classifies masses, while Mask R-CNN identifies tumor borders and sizes, crucial for staging cancer. Trained on CBIS-DDSM dataset, BNS datasets In breast dataset, this combined approach reduces False Positive Rate (0.049%) and False Negative Rate (0.029%) and achieves a high Matthews correlation coefficient (92.02%), outperforming the baseline YOLOv5. This approach improves early diagnosis accuracy, aiding clinicians in better prognosis and treatment planning. The author [22] presents a deep learning-based ensemble classifier for early and precise breast cancer detection. Integrating transfer learning models with advanced techniques like residual learning and skip connections achieves high accuracy. Results on various datasets show significant performance: 99.17% accuracy for abnormality detection and 97.75% for malignancy on mini-DDSM, 96.92% and 94.62% on BUSI ultrasound, and 97.50% on BUS2 ultrasound. This approach proves versatile across multimodal datasets, promising improved diagnostic capabilities in clinical settings.

The author [23] underscores the importance of early breast cancer detection using AI and machine learning advancements. Their study focuses on improving diagnostic accuracy for IDC and DCIS subtypes from mammograms, employing the CNN Improvements for Breast Cancer Classification (CNNI-BCC) model. This research introduces an efficient deep learning approach validated with a dataset of 3002 images, demonstrating high accuracy with reduced computational demands across varied mammogram densities. The author [24] employed early detection of breast cancer is critical for saving lives, and researchers are exploring new methods like thermal imaging for accurate diagnosis. Recent advances in deep learning have led to the development of lighter models, such as using SqueezeNet 1.1, fine-tuned on thermal images. These models incorporate transfer learning and feature selection techniques, alongside hybrid optimization strategies like Genetic Algorithm and Grey Wolf Optimizer. This approach reduces computational complexity while achieving high accuracy, showing potential for precise identification of malignant and healthy breast conditions with minimal computational resources.

The author [25] proposes using Fuzzy C Means segmentation and an IWCA-APSO-based Ensemble Extreme Learning Machine model for breast cancer detection. Validated on the INbreast dataset, the approach achieves high sensitivity (99.67%), specificity (99.71%), and accuracy (99.36%), with a computational time of 23.8751 seconds. These results indicate significant improvement over traditional methods in breast cancer classification. The author [26] proposed ensemble learning model for breast cancer detection. Having 317880 samples which split in 80% and 20% for the training and validation process respectively. They got 91.33% accuracy for detection the breast cancer. Table 1 shows the previous state-of-the-art along with techniques been used, dataset and result of study.

**Table 1.** Previous study of breast cancer.

| Author | Technique | Dataset | Result |
| --- | --- | --- | --- |
| [15] | YoLoV5 | 307 lesions | Map 0.621 |
| [16] | RF+SVM | - | Accuracy 99.99% |
| [17] | SDM-WHO-RNN | - | Accuracy 97.90% |
| [18] | MWI phase imaging | - | Sensitivity 90.9% |
| [19] | Transfer learning + LSTM | 780 images | F1 score 99.0% |
| [20] | ResNet50 | 986 images | Accuracy 98.98% |
| [21] | YOLOv5 | 2120 images | MCC 92.02% |
| [22] | AlexNet + MobileNet + Res-Net | 9684 images | Accuracy 97.50% |
| [23] | CNNI Model | 3002 images | Accuracy 96.49% |
| [25] | Ensemble extreme learning model | 410 mammograms | Accuracy 99.36% |
| [26] | Ensemble learning model | 317880 images | Accuracy 91.33% |
| Proposed | Few-Shot-Meta-Learning | Total 200 images, with 1-shot and 5-shot | Accuracy approx. 96.00% for 5-shot |

### Preliminary Concepts

Before providing an in-depth analysis of our framework, we will start by outlining key principles of few-shot classification. This foundational step is essential as it may be new to some readers.

### Supervised Classification

In this classification, we work with a dataset “D” that includes both training data D_train_ and testing data D_test_. Training dataset, D_train_ is composed of labeled instances 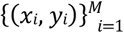 where M is the number of training examples, and y_i_ belongs to the set {1,…,A_total_}. Here, A_total_ indicates the total number of categories within the training data. Our task is to develop a model f_θ_(x), with θ representing the model parameters, trained on D_train_. This model should be capable of predicting the label *ŷ* ∈ {1,…,A_total_} for unknown sample x_p_ in test set D_test_, which consists of 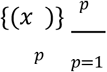.

### Few-shot Classification

Instead of using the traditional method, we utilize a meta-dataset composed of three distinct subsets: D={D_base_,D_val_,D_novel_}. Importantly, the category labels, denoted as CC, are chosen such that CC_base_, CC_val_ and CC_novel_ have no overlap in categories. Our ultimate goal is to develop a model, MD, using the dataset Dbase. This model needs to be proficient at quickly adapting to new categories presented in the Dnovel dataset, even when only a few support samples (typically 1 to 5 examples per category) are available. We will use the Dval dataset exclusively for hyperparameter tuning and for identifying the best-performing model.

Following the Few-Shot Learning (FSL) protocol as described in references [27][28], models are generally evaluated through a series of Tasks. These Tasks involve the classification of K examples among N classes and are known as D_T_ or episodes. Each episode is made up of two sets: a support set SS_i_ and query set QS_i_. The support set SS_i_ has N unique categories, each with P labeled samples. Thus, the support set SS_i_ contains a total of N * P labeled samples, which are used for training. Query set QS_i_, however, includes the same N categories but contains Q unlabeled samples that need been classified.

Episodes are constructed in the same manner for both training and testing phases. To illustrate, consider a 2-way 1-shot classification task during testing. In this case, the training episode would consist of N = 2 categories, each with P = 1 labeled sample. Figure 1 visually depicts a 2-way 1-shot episode. It is essential to understand that, within the Few-Shot Learning (FSL) framework, in this approach, an entire task or episode is considered a single training instance, marking a notable departure from traditional machine learning methods.

**Figure 1.**
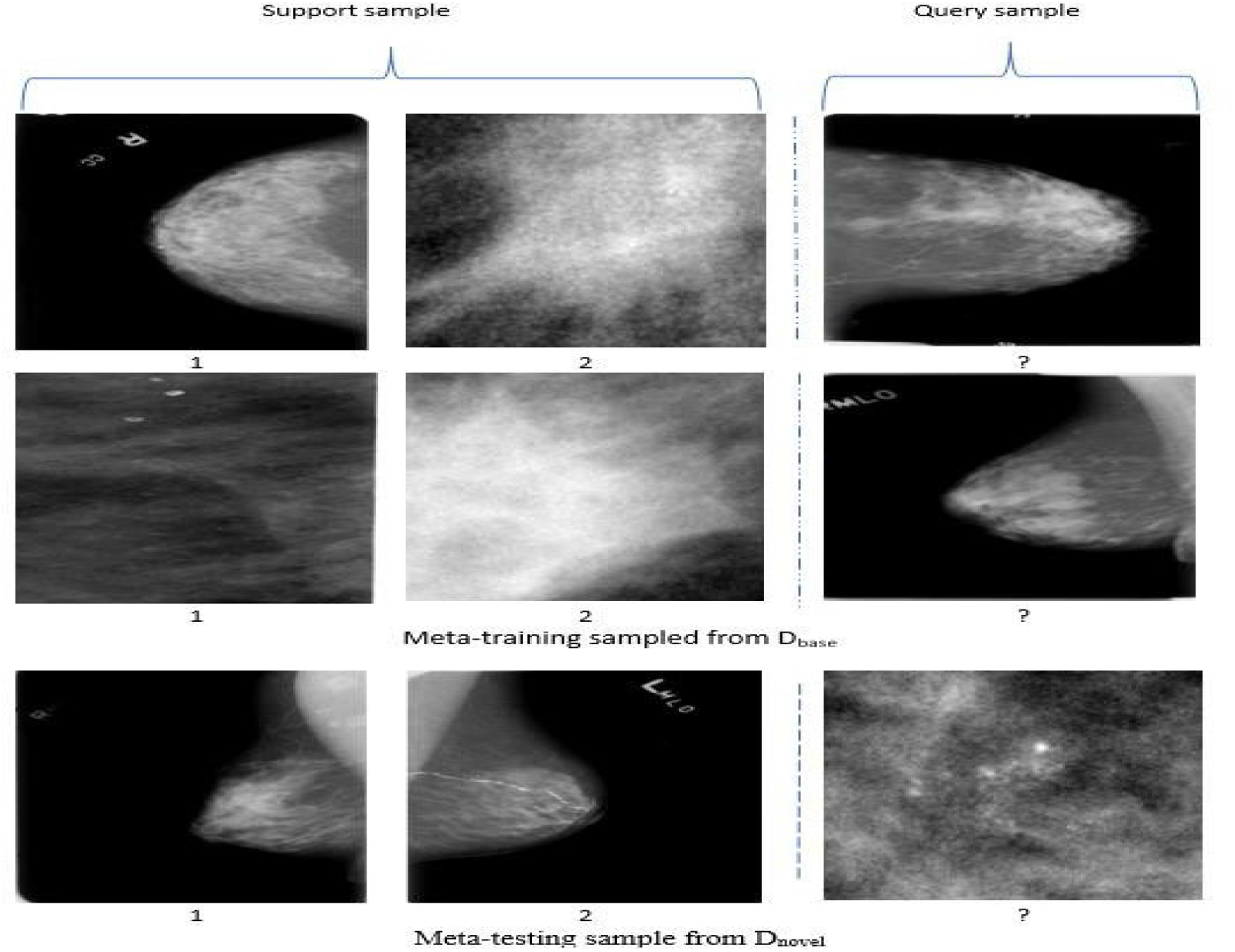
Depiction of meta-training and meta-testing sets for 2-way 1-shot classification tasks.

### Proposed Research Study

#### Comprehensive Framework

This study introduces a tailored meta-learning method designed to diagnose the breast cancer, effectively tackling data scarcity issues. Our methodology is structured around five crucial stages: preprocessing, segmentation, feature extraction, meta-training, and meta-testing, as illustrated in Figure 2. Initially, we undertook image preprocessing to improve both the quality and clarity of the images. For segmentation, U-Net is being applied. Subsequently, we developed a feature extractor using a foundational dataset D_*base*_ to generate a strong representation of mango crop images. We utilized a conventional classifier, training it across all categories within the base dataset by minimizing the cross-entropy loss. By detach the final fully connected layer, we obtained 512-dimensional feature representation, referred to as *f*_*θ*_.

**Figure 2.**
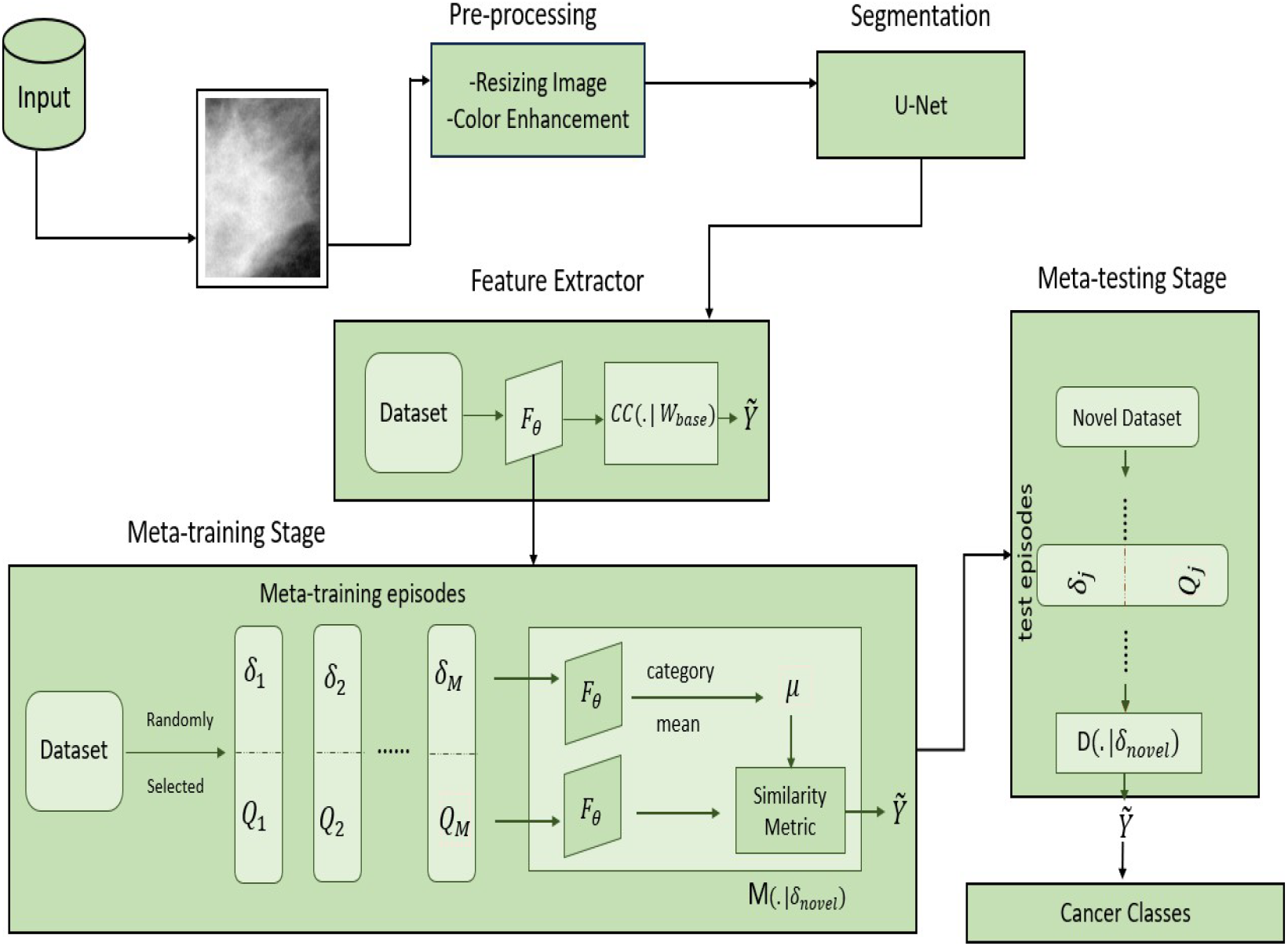
Proposed Breast Cancer Detection Framework.

During the meta-training stage, the meta-learning classifier M was trained across multiple episodes without freezing the feature extractor *f*_*θ*_. Instead, we optimized the feature extractor directly to reduce generalization errors across episodes. This approach enabled effective classification of breast cancer stages using few-shot learning techniques. In each episode, we compared the features of query images with the mean features of support images using a scaled cosine distance metric. Goal of meta-training was to minimizing the N-way classification loss on the query set. During meta-testing stage, we assessed the performance of the meta-learning classifier M by using episodes sampled from a new dataset D_*novel*_. This phase evaluated the classifier’s ability to adapt to new and unseen categories of breast cancer, analogous to the meta-test set.

### Proposed Few-shot meta learning approach

Meta-learning represents a cutting-edge machine-learning technique that allows models to learn the process of learning, making it particularly advantageous for scenarios with limited or noisy data akin to the complexities found in breast cancer detection. In our research, we have developed a new meta-learning strategy specifically designed to detect breast cancer. We assessed the performance of this approach using a diverse dataset that includes ultrasound images representing different stages of breast tissue characteristics and abnormalities.

### Dataset and Pre-processing

In this research dataset, a collection of 100 images depicting detection of breast cancer was compiled from sources such as medical databases [29]. These images were classified into two distinct categories: those showing malignant and benign tumors. All images were standardized to a resolution of 256*256 pixels. To guarantee the robustness of our model, we meticulously followed industry best practices during the preprocessing phase. We ensured that our dataset was diverse, maintained high resolution, and adhered to appropriate resizing and formatting standards. This careful preparation aimed to improve the overall quality and representativeness of the data, crucial for effective model training.

The preprocessing steps were designed to enhance data quality, reduce noise, and streamline analysis. For this study, we applied several key preprocessing techniques to mammogram images. Notably, we used histogram equalization to enhance image contrast, making potential breast cancer markers more visible. This step was essential to improve feature detection and ensure the reliability of our model’s predictions.

To improve the contrast and clarity of mammogram images for breast cancer detection, we used histogram equalization as a preprocessing technique. This method enhances the visibility of important features by redistributing the pixel intensity values, which broadens the dynamic range of the images. Consequently, critical details that may indicate the presence of breast cancer become more discernible. This enhancement of image quality is a crucial step in ensuring the accurate identification of potential breast cancer markers.

### U-Net

Biomedical images, such as those used for detecting breast cancer, often display intricate patterns and variable edges that can complicate segmentation. To tackle this issue, the author [30] proposed an architecture for skin segmentation that combines high-level features from a deep decoding layer with low-level features from a shallow encoding layer, achieving detailed segmentations. This method has demonstrated efficacy not only in natural images but also in biomedical applications [31]. Expanding upon this idea, the author [32] introduced the U-Net architecture, which utilizes skip connections to improve cell tracking in biomedical imaging.

Our network architecture for breast cancer detection, inspired by U-Net, consists of a dual-path structure: an encoding path for down sampling and a decoding path for upsampling, as shown in Figure 3. The encoding path includes five convolutional blocks, each comprising two 3*3 convolutional layers with a stride of 1 and rectifier activation functions. These blocks progressively increase the number of feature maps from 1 to 1024. Spatial dimensions are reduced using 2*2 max pooling at the end of each block, except the last one, thereby decreasing the feature map sizes from 240*240 to 15*15.

**Figure 3.**
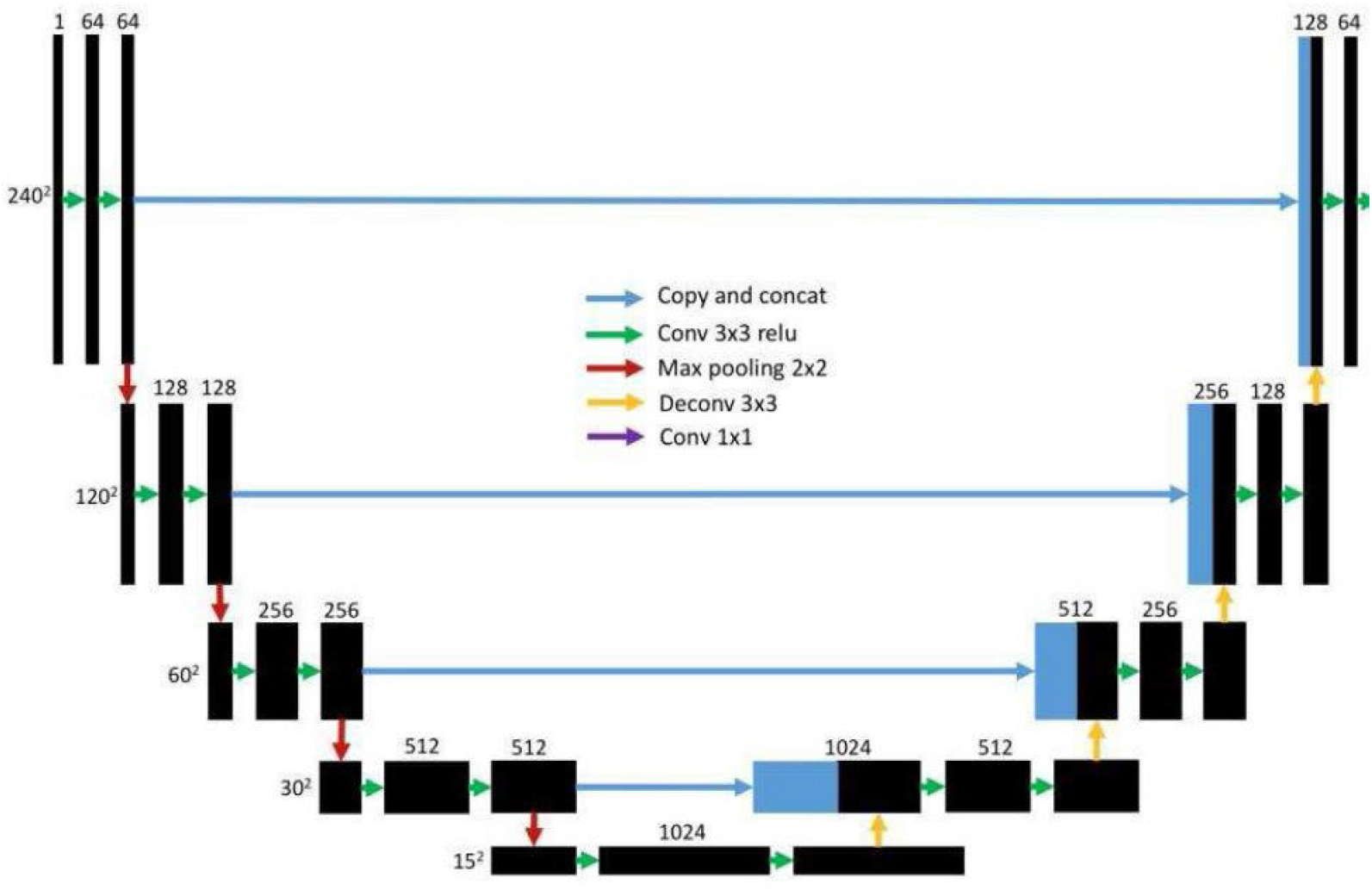
U-Net Architecture [33]

In decoding, each block begins with deconvolutional layer featuring 3*3 filter size and a stride of 2*2. This operation doubles the spatial dimensions of the feature maps while halving the number of feature maps, effectively increasing their size from 15*15 to 240*240. This approach allows our network to efficiently reconstruct high-resolution images from the compressed features, facilitating precise breast cancer detection.

In our design, each up-sampling block incorporates two convolutional layers to decrease the number of feature maps. These layers operate on the combined output of the deconvolutional feature maps and the feature maps from the corresponding encoding block. To diverge from the original U-Net structure, employ zero padding in all convolutional layers within both the downsampling and up-sampling paths, ensuring the output dimensions remain unchanged.

In the final stage of the network architecture, a crucial step involves the application of a 1*1 convolutional layer. This layer plays a pivotal role by effectively reducing the dimensionality of the feature maps to just two channels. These channels specifically cater to delineating between foreground and background segments within the image. Notably, the design circumvents the use of fully connected layers, adhering to a streamlined and efficient computational approach.

### Extraction of features

In our study focused on breast cancer detection, we devised a feature extractor labeled *f*_*θ*_, which operates with adjustable parameters denoted as ‘θ’. This extractor is designed to transform input data originating from our primary dataset, referred to as D_*base*_, into a streamlined 512-dimensional feature vector optimized for comprehensive comparisons. Our approach leveraged the robust architecture of DenseNet-121 as the foundational framework for training a classifier across all relevant categories. A key innovation of our methodology involves customizing this network architecture into a feature extractor by excluding its final dense (fully connected) layer. This strategic modification enhances our ability to discern nuanced features crucial for accurate breast cancer detection. Throughout our research, we also explored alternative backbone architectures, ensuring a meticulous evaluation of the most effective model configurations for our specific application.

To ensure uniform processing of images from our dataset D_*base*_ in our breast cancer detection study, we implemented a preprocessing step where all images were resized to a standardized dimension of 80*80 pixels. This preprocessing step was crucial for maintaining consistency in input dimensions across our entire dataset.

Our chosen model architecture, DenseNet-121, depicted in Figure 4, features a series of dense blocks interspersed with transition layers. Each dense block comprises consecutive 3*3 kernel convolutional layers, bolstered by Batch Normalization (BN) and Rectified Linear Unit (ReLU) activation functions. Configuration of these convolutional layer varies across the dense blocks, characterized by channel configurations {C_1_, *C*_2_, *C*_3_, …, *C*_*n*_}, where n denotes total number of dense block in our model. Specifically, in our DenseNet-121 setup, these values are tailored to 64, 128, 256, and 512 channels for the respective dense blocks.

**Figure 4.**
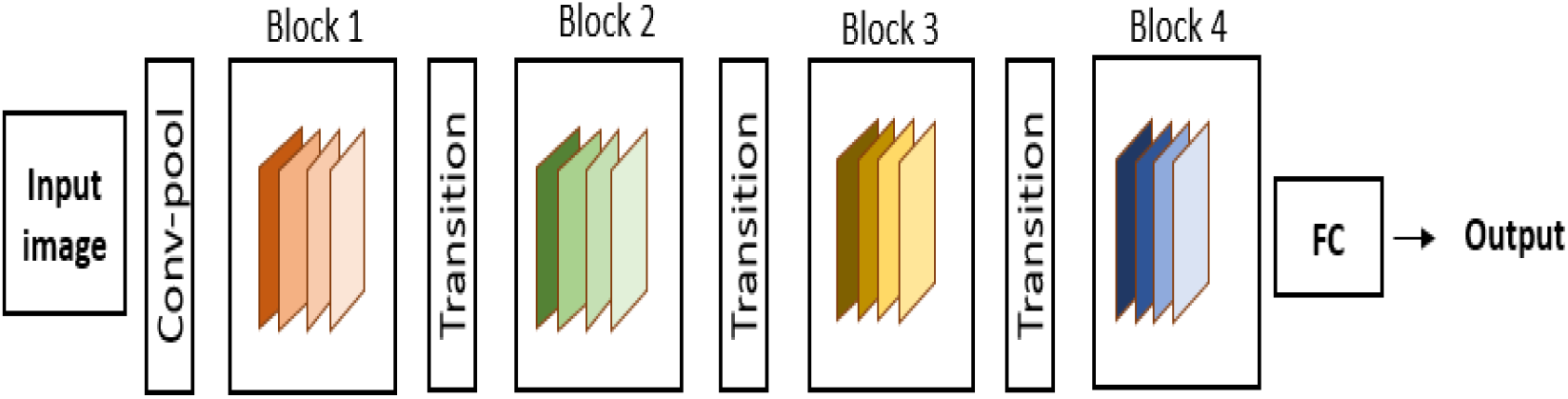
Architecture of DenseNet-121

This architecture design not only ensures efficient feature extraction from input images but also optimizes the model’s ability to discern intricate patterns relevant to breast cancer detection. By leveraging these structured layers and channel configurations, our approach aims to enhance both the accuracy and reliability of our diagnostic outcomes.

After each dense block, our architecture incorporates a transition layer comprising a 1*1 convolutional operation, Batch Normalization, ReLU activation, and 2*2 average pooling. These layers effectively reduce spatial dimensions and filter complexity while preserving essential feature information for subsequent processing stages in our breast cancer detection model. At the culmination of feature extraction, a 5*5 global average pooling layer condensed DenseNet’s feature maps into a compact 512-dimensional representation, pivotal for subsequent tasks in our breast cancer detection approach.

### Meta-Training Stage

Meta-learning objectives improve breast cancer diagnosis efficiency by leveraging insights from various diagnostic tasks (episodes). In an N-way K-shot setup, we train a meta-learning model F(· | S) to minimize prediction error using episodes drawn from the training data, each with K samples per category. This results in N * P samples for training and N * Q samples for testing. Despite limited support samples, the M classifier’s parameters are shared across episodes, reducing the need for extensive training examples. An additional meta-validation set fine-tunes the model’s hyperparameters. Figure 5 illustrates our meta-learning workflow.

**Figure 5.**
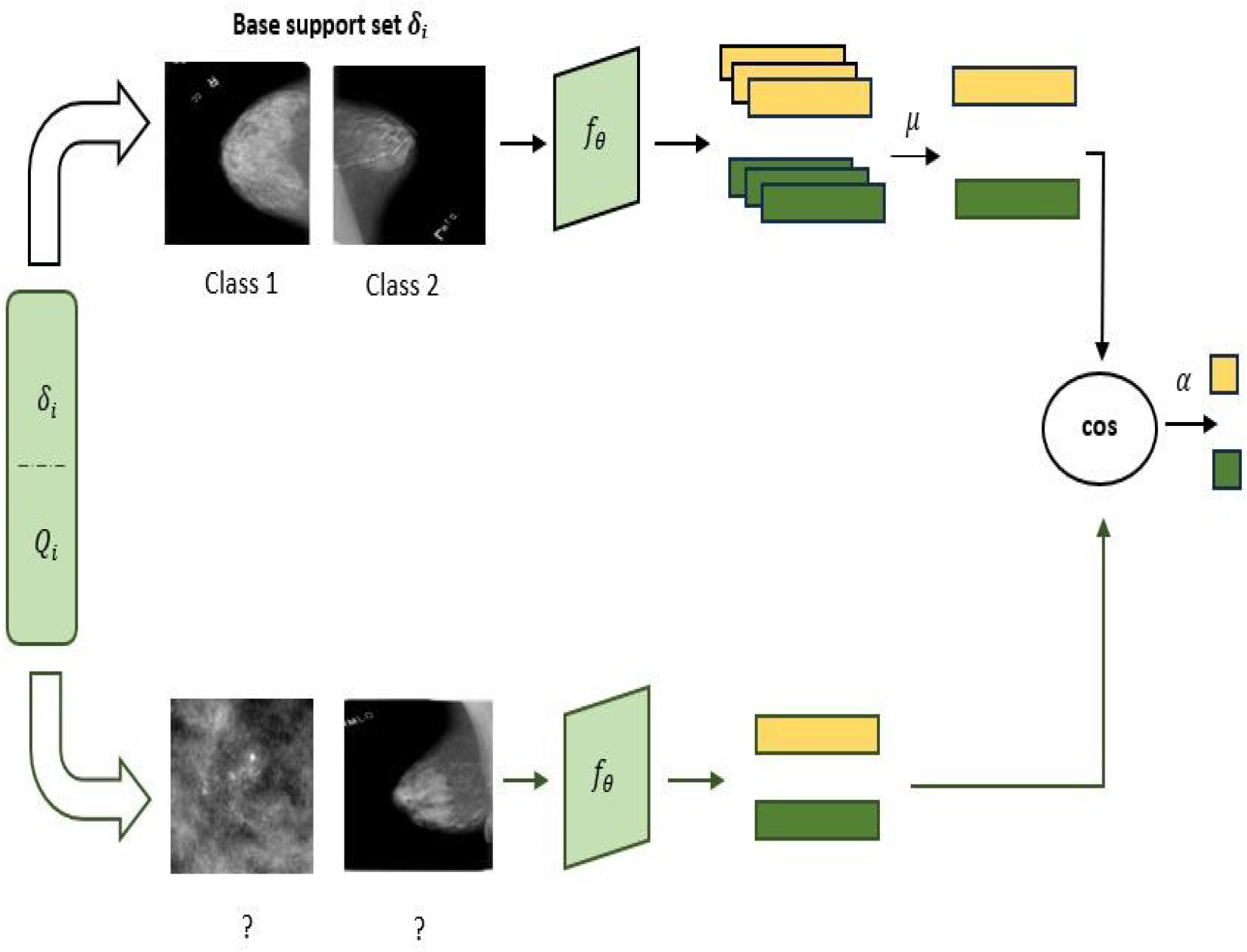
Developing meta-training for addressing 2-way 1-shot classification

Within an episode where we have a support-set S, S_c_ represents a subset of S that includes all samples belonging to category c. The prototype *ω*_*c*_ is derived as the average vector of embeddings of Sc. These embeddings are obtained using a pre-trained feature extractor denoted as *fθ*, which involves learnable parameters θ in detailed in section 4.2.

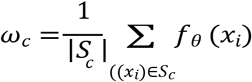

To determine the probability that q query sample x belongs to category c, we assess the similarity between its feature embedding *f*_*θ*_ (x) and the centroid *ω*_*c*_ of that category. By employing cosine similarity as a distance metric, the prediction is formulated as below:

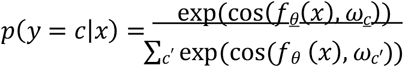

Here, cos(*f*_*θ*_(*x*), *ω*_*c*_) denotes the cosine similarity between the feature embedding *f*_*θ*_ (x) and the centroid *ω*_*c*._ It measures the directional alignment between these two vectors as an indicator of their similarity.

Drawing from insights in [34], trainable scalar-parameter α to adjust the initial range of cosine similarity been introduced, which conventionally spans from -1 to 1. In our experiments, we initialize α to 10. This scaling of the similarity metric enhances its suitability for integration into the subsequent softmax layer. As a result, the formulation of the predictive probability is as follows:

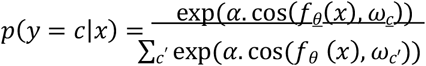

### Meta-Testing Stage

We train the neta-learning model F(· | *SS*_*base*_) on the base dataset SS_base_, and then evaluate its generalization capability using a separate dataset, D_novel_. It is important to note that the categories in D_novel_ are completely new and were not encountered in course of meta-training phase.

In meta-testing step, new episodes frawn from D_novel_, commonly referred to as the meta-test and denoted as 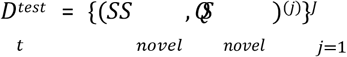. The trained model then adapts to these unseen categories by utilizing the new support set SS_novel_ to make predictions.

## Results

This section This section presents a detailed overview of our implementation and a description of the dataset utilized in our study.

### Dataset Description

In the existing dataset, there were a total of 10,239 images. However, the approach we are applying, few-shot meta-learning, is designed to work effectively with a smaller number of samples. Therefore, we have selected 100 samples for each class. The details are presented in the table 2 below.

**Table 2.**
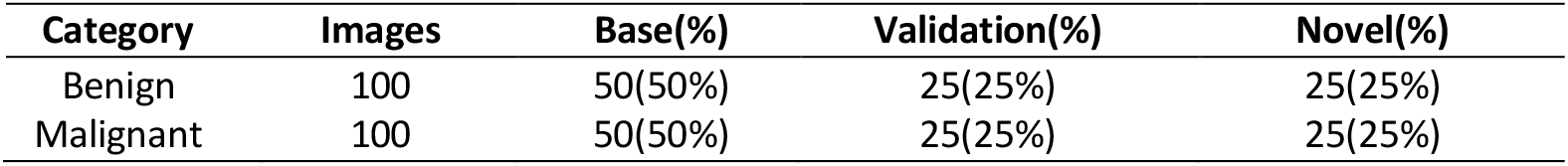
Dataset Overview.

| Category | Images | Base(%) | Validation(%) | Novel(%) |
| --- | --- | --- | --- | --- |
| Benign | 100 | 50(50%) | 25(25%) | 25(25%) |
| Malignant | 100 | 50(50%) | 25(25%) | 25(25%) |

### Detailed implementation overview

Inspired by Vinyals et al. [35], we implemented an experimental framework for N-way classification with K shot, where N = 2 and K = 1. During meta-training, we generated few-shot training batches using episodes. Each episode randomly selected 2 categories from the database Dbase. Our setup included 4 episodes per batch, with batch sizes adjusted to fit GPU memory. In each training episode, the support set matched the requirements for meta-testing. For instance, for a 2-way 1-shot classification in testing, training episodes were set with N = 2 and K = 1.

During meta-training, each category had K query samples, while in meta-testing, there were 15 query samples per category. In the realm of few-shot learning, we vary our training approach by randomly selecting episodes. Despite each episode containing a small subset of training samples, employing a large number of episodes—such as the 1000 utilized per epoch—ensures thorough coverage of the dataset throughout training.

We utilized DenseNet-121 as our core architecture, modifying it by excluding the fully connected layer to yield 512-dimensional feature vectors per input. The training process was managed using Stochastic Gradient Descent (SGD) with a momentum parameter set to 0.9. The initial learning rate was 0.1, which decayed by a factor of 0.1 over time. The feature extractor was trained for 100 epochs with a batch size of 128, distributed across 4 GPUs. Additionally, we applied a weight decay of 0.0005 to refine the DenseNet-121 model. Computational tasks were executed within a Jupyter Notebook environment, leveraging the computational power of an NVIDIA GeForce GTX Titan X GPU (RTX 2080 Ti).

### Outcomes and Analysis

To evaluate our method’s effectiveness, we conducted experiments using the standard few-shot classification setups: 2-way 1-shot and 2-way 5-shot. In the 2-way 1-shot setup, one support sample per category is used for testing, while in the 5-shot setup, five support samples per category are used. Each episode was assessed with 15 query images per category, and we calculated the mean accuracy over 2000 random episodes from the novel set. Employing a more advanced model such as DenseNet-121, rather than the initial Conv-4 used in Model-Agnostic Meta-Learning (MAML), has the potential to enhance performance. Additionally, we explored the efficacy of the traditional classification algorithm, D-CNN, in contexts with limited data availability.

Deep Convolutional Neural Network is model after the biological architecture of the animal visual system, especially the visual cortex. This approach mimics the behavior of specialized cells in the cortex that respond to particular areas, or receptive fields, of the visual field. In traditional fully connected neural networks, each neuron is linked to every neuron in the preceding layer, a D-CNN neuron is connected only to a specific receptive field in the layer before it.

A typical D-CNN includes three essential layers: the Convolutional Layer, the Pooling Layer, and the Fully Connected Layer [36][37]. Model-Agnostic Meta-Learning (MAML) is a few-shot learning method that prepares a model to adapt rapidly to new tasks with minimal data. During meta-training, MAML initializes the model on a source task and then trains it across a variety of tasks, allowing the model to efficiently adjust its parameters. This results in strong performance on novel, unseen tasks with limited examples. MAML is especially valuable when collecting extensive labeled data for each task is impractical, making it a powerful tool for scenarios with scarce data [38].

Both 1-shot and 5-shot settings, the proposed approach outperforms MAML with DenseNet-121 across both datasets. Conversely, D-CNN shows poor performance in these scenarios because it is not designed for few-shot classification. Traditional CNNs tend to overfit when trained on very limited data, whereas meta-learning methods yield better results. Table 3 provides a comparison of the results for MAML, D-CNN, and the proposed method using the DenseNet-121 backbone.

**Table 3.** Analysis of Few-Shot Classification Dataset.

| **Method** | **Backbone** | **1-shot** | **5-shot** |
| --- | --- | --- | --- |
| MAML | DenseNet-121 | 61.43 ± 0.85 | 75.93 ± 0.68 |
| D-CNN | DenseNet-121 | 45.00 ± 5.32 | 62.71 ± 4.30 |
| Ours | DenseNet-121 | 71.10 ± 0.21 | 96.10 ± 0.13 |

The comparison of MAML, D-CNN, and our DenseNet-121-based method across our datasets is depicted in Figure 6, where patterned bars represent results with 95% confidence intervals. Our method consistently outperforms both MAML and D-CNN, demonstrating superior performance across all evaluations.

**Figure 6.**
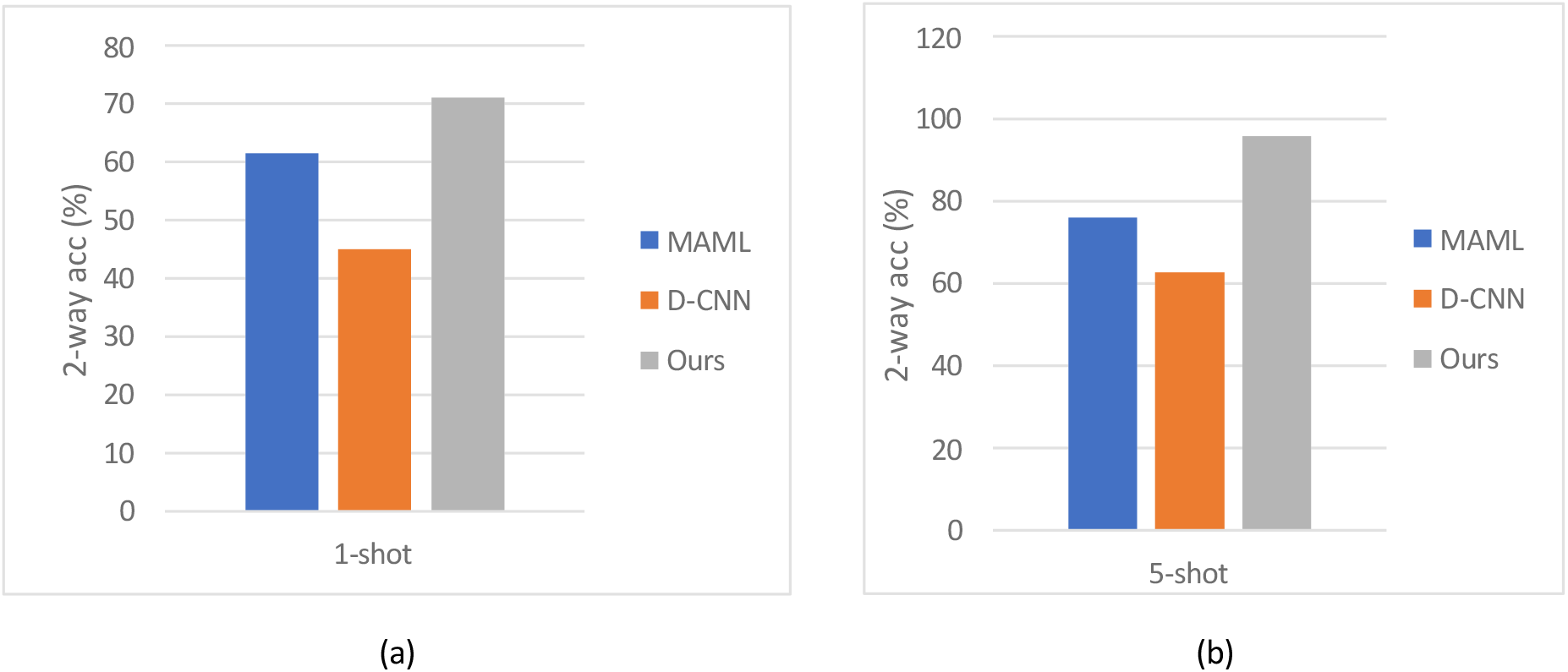
Illustrates few-shot classification results on our dataset using DenseNet-121, with 95% confidence intervals shown for both 1-shot (a) and 5-shot (b) scenarios.

Utilizing a more efficient variant of first-order MAML, known to approach the performance of the full version, we enhance classification accuracy through class center calculations and a cosine distance metric with adjustable scaling. In Section 5.4 of our study, an ablation experiment examines the impact of metric selection on outcomes. Drawing from insights in [39], which highlight the importance of task-specific feature extraction, we introduce a dynamic feature extractor optimized with a designated support set labeled ‘SS’. To streamline complexity, an additional logit head facilitates auxiliary co-training. In contrast, our approach focuses on training a standard M-way classifier on established categories, omitting extra parameters. Instead, the encoder *f*_*θ*_ benefits from refinement by removing the final FC layer, leveraging its weights for enhanced meta-training.

Throughout the analysis depicted in Figure 7, an intriguing pattern emerges: our model initially demonstrates improved generalization across base categories within the first 90 epochs. However, this trend contrasts sharply with its ability to generalize to novel categories, which shows a decline. This decrease in test performance is attributed to potential overfitting, likely stemming from limited supervised data availability. Further investigation into this phenomenon is detailed in Section 5.4.

**Figure 7:**
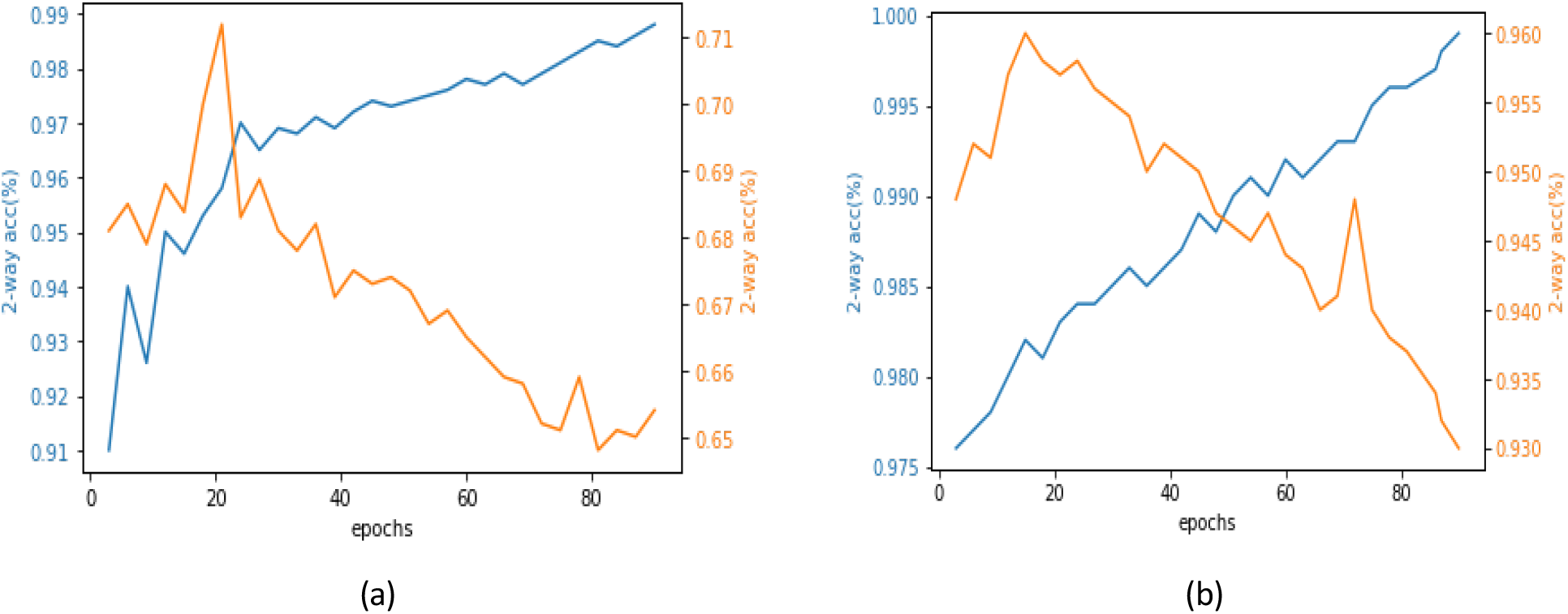
Generalization differences during the meta-learning stage, with base generalization shown in blue and novel generalization in orange.

### Analytical Findings

#### Dataset Size implications

To evaluate the influence of dataset size, we created a reduced version of our complete dataset, which originally contains 100 images per category. This smaller version, termed the “mini dataset,” includes only 50 images per category. Table 4 presents the 2-way 1-shot and 2-way 5-shot accuracies for both the original and mini datasets. By dividing the dataset in this manner, The study focused on examining how different dataset sizes influence the model’s performance and evaluating the robustness of the approach. The findings show that the larger dataset consistently outperforms the smaller one, achieving 6.86% and 5.78% higher accuracy in the 2-way 1-shot and 2-way 5-shot scenarios, respectively.

**Table 4.**
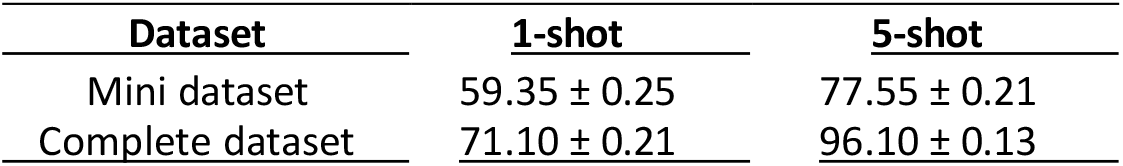
Performance comparison: Mini vs. Full Dataset.

| <u>Dataset</u> | <u>1-shot</u> | <u>5-shot</u> |
| --- | --- | --- |
| Mini dataset | $59.35 \pm 0.25$ | $77.55 \pm 0.21$ |
| Complete dataset | $71.10 \pm 0.21$ | $96.10 \pm 0.13$ |

Section 5.3 highlights a consistent observation from our experiments, depicted in Figure 8. Initially, our model achieves peak accuracy within the first 40 epochs. To delve deeper into the issue of generalization, we examined generalization curves for different shot counts on the dataset. These curves, shown in Figure 8, distinguish between novel generalization (orange) and base generalization (blue). Intriguingly, we observed a similar trend: while the model performs well on unseen data from the base set, its performance on novel tasks declines. This suggests that simply increasing the number of labeled support instances does not solely account for the decrease in test performance; instead, the discrepancy likely arises from differing objectives between the novel and base sets. During meta-training, our model may become overly specialized to the base set, adversely affecting its performance on novel tasks. Addressing this generalization gap in few-shot learning presents a significant challenge. Future investigations focusing on regularization techniques could potentially mitigate this gap.

**Figure 8.**
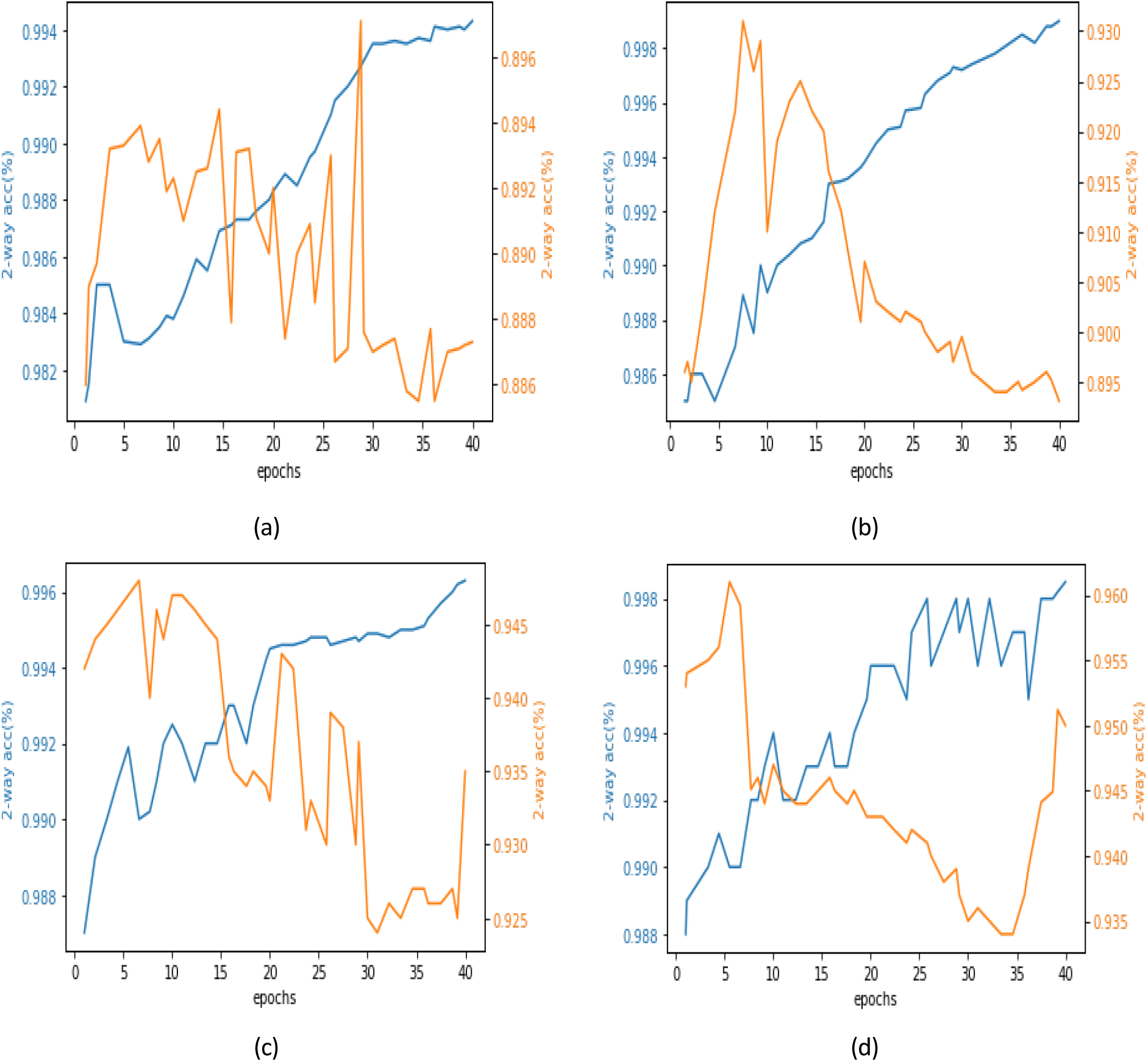
Illustrates the performance variability, accompanied by 95% confidence intervals, for 5-way classification using the DenseNet-121 backbone. The analysis considers different numbers of shots applied to the dataset for 10-shot, 20-shot,30-shot and 40-shot in (a),(b),(c),and (d) respectively.

Table 5 summarizes the performance of several techniques for the task at hand. Ali et al.[42] utilized a CNN with impressive results, achieving 90.00% accuracy, demonstrating the strength of convolutional neural networks in this context. Wu et al. [40] applied SVM, also achieving 90.00% accuracy, showcasing the effectiveness of support vector machines. G Isik et al. [41] explored few-shot learning, achieving 83.1% accuracy, highlighting its potential in scenarios with limited labeled data. Our approach, leveraging meta-learning with a DenseNet-121 backbone, achieved competitive performance with an accuracy of 96.10% ± 0.13. This comparison underscores the versatility of different methodologies and their ability to address challenges in machine learning tasks effectively.

**Table 5.** Comparison of proposed study with previous state-of-the-art.

| Approach | Technique | Accuracy |
| --- | --- | --- |
| [42] | CNN | 90.00% |
| [41] | SVM | 90.00% |
| [40] | Few-shot | 83.10% |
| Ours | Meta-learning with DenseNet-121 backbone | 96.10 ± 0.13 |

## Conclusions

The recent surge in few-shot learning has piqued our interest in exploring its application to breast cancer detection. We demonstrate the efficacy of few-shot learning in the context of classifying breast cancer subtypes, revealing valuable insights from limited examples. Our meta-learning framework, utilizing DenseNet-121, effectively generalizes to new cancer subtypes with minimal samples. Through optimizing the classifier with cosine distance and a scalable parameter during meta-training, we achieved promising results: approximately 71% classification accuracy for new cancer subtypes with a single sample, and about 96% accuracy with five support samples. Furthermore, our exploration of dataset sizes, various metrics, and the number of support shots uncovers potential challenges in generalization within the meta-learning framework. Continued research in this domain holds promise for improving future diagnostic accuracy and treatment strategies in breast cancer.

## Data Availability

The data sets are available upon request.

## Author Contributions

Conceptualization, N.U. and A.B.; methodology, N.U. and A.B.; software, N.U. and U.S.; validation, N.U., U.S., H-S. C.and A.B.; formal analysis M.H.A. and H-S. C., writing— original draft preparation, N.U.; writing—review and editing, A.B., M.H.A., and H-S. C.; visualization, U.S. All authors have read and agreed to the published version of the manuscript.

## Data Availability Statement

Data can be available on request.

